# The response to BTN62b2 booster doses demonstrates that serum antibodies do not predict the establishment of immune B-cell memory in common variable immune deficiencies

**DOI:** 10.1101/2022.12.24.22283775

**Authors:** E. Piano Mortari, F. Pulvirenti, V. Marcellini, S. Terreri, A. Fernandez Salinas, S. Ferrari, G. Di Napoli, D. Guadagnolo, E. Sculco, C. Albano, M. Guercio, S. Di Cecca, C. Milito, G. Garzi, A.M. Pesce, L. Bonanni, M. Sinibaldi, S. Di Cecilia, C. Agrati, C. Quintarelli, S. Zaffina, F. Locatelli, R. Carsetti, I. Quinti

## Abstract

In patients with common variable immune deficiencies, primary vaccination followed by two booster doses is recommended for protection against COVID-19. Seroconversion has been shown in 60% of patients. We have no information on whether serum antibodies reflect the generation of durable immune memory.

In a longitudinal study on 47 common variable immune deficiencies patients who received the third and fourth vaccine dose, we show that the measurement of specific antibodies is not sufficient to predict the establishment of immune memory and the ability to respond to antigen re-exposure.

Our results indicate that the combination of antibodies and memory B cells responses represents a more reliable read-out of vaccine immune efficacy in vulnerable patients.

This analysis may not only identify individuals remaining unprotected after vaccination and unable to respond to additional booster doses, but also address the search for the underlying immune defect and suggest patient-tailored management strategies.

---

Infection prophylaxis by vaccine administration with inactivated or non-viable vaccines is a current practice in patients with Inborn Errors of Immunity (IEIs)^1, 2^. Furthermore, the impaired response to immunization is also included in the diagnostic criteria of many IEIs such as Common Variable Immune Deficiencies (CVIDs)^1, 3, 4^, a complex and heterogeneous group of IEIs characterized by hypogammaglobulinemia and impaired antibody production^1, 2^. Due to the heterogeneity of CVIDs, humoral immune responses to immunization range from partial to absent antibody production and the grade of impairment correlates with prognosis^5, 6^. The antibody defect is treated by immunoglobulin replacement therapy (IgRT) containing a wide spectrum of antibodies, including those directed to vaccine antigens, with the aim of reducing the number and severity of infections^7^. Diagnostic evaluations of humoral responses to vaccines may not be informative in patients with IgRT^8^. To overcome this limitation, the analysis of antibody responses to neoantigens has been proposed^9^, but rarely used in clinical practice.

Over the last two years, the SARS-CoV-2 pandemic has led to the rapid development of effective vaccines against the viral spike protein, an antigen that humans have never encountered before. Vaccination against this novel antigen has offered the opportunity to study the response to a primary immunization in wide cohorts of patients with IEIs^10, 11^, including CVIDs^11–23^.

Published data indicate that after two vaccine doses, 60% of CVIDs patients are able to seroconvert^10^ with a lower magnitude of antibody response and a reduced virus-neutralizing function compared to control ^12–14, 16, 19^.

We have no information, however, on whether the presence of serum antibodies reflects the establishment of immune memory able to be re-activated by booster vaccination or infection. In immunocompetent subjects, antibodies are just the tip of the iceberg of the response to immunization. Persistent memory T and B cells (MBCs) are indispensable for the protective response in case of re-encounter with the pathogen ^24, 25^. We have shown that vaccine-induced antibodies and MBCs have divergent kinetics in healthy subjects: whereas antibody titers are high early after immunization and rapidly decline, MBCs persist and increase in time^25^. In case of breakthrough infection or administration of booster vaccine doses, MBCs rapidly react, increasing in numbers and producing antibodies^25^. Similarly, memory T cells frequencies and absolute numbers do not decline in time ^26^.

MBCs include IgM and switched MBCs. IgM MBCs are generated by a T- and germinal center (GC)-independent mechanism, carry few somatic mutations^27–33^ and serve as first-line protection against infection^34^. Switched MBCs are generated in the GC with the indispensable help of T cells expressing the CD40 ligand^33, 35^. MBCs accumulate somatic mutations in the GC and are selected for their increased affinity to the stimulating antigen^36^.

In control individuals, IgM MBCs with low affinity for the viral spike protein serve as substrates for the generation of high-affinity MBCs that are class-switched and are mainly of the IgG isotype^25, 37^. Recently, it has been suggested that antibodies produced by IgM MBCs may play an important role in the defense against SARS-CoV-2, as demonstrated by the potent neutralizing activity of IgM monoclonal antibodies cloned from MBCs of individual convalescent from COVID-19^38^.

In this longitudinal study carried out in CVIDs patients naïve to SARS-CoV-2 infection who completed the primary cycle with the BNT162b2 mRNA vaccine followed by one (3^rd^ dose) or two (4^th^ dose) booster doses, we combine the data on the production of specific antibodies and MBCs. Using this approach, we demonstrate that serum antibody measurement is not sufficient to predict the establishment of immune memory in CVIDs patients. We also show that the performance of the immune system in the response to vaccination in vivo may be used to discriminate patients with different mechanisms at the base of the individual immune defect and suggest patient-tailored management strategies.

## Results

### Patients

Forty-seven CVIDs patients (median age 53.5 years (IQR 45.7-66), females 28 (58.3%)) immunized with the 3^rd^ dose of the mRNA BNT162b2 vaccine were included in the analysis. Six months apart, the 4^th^ dose was administered to 25/47 patients, because 12 participants were infected by SARS-CoV-2 and 10 patients refused the additional booster doses (Figure 1).

**Figure 1.**
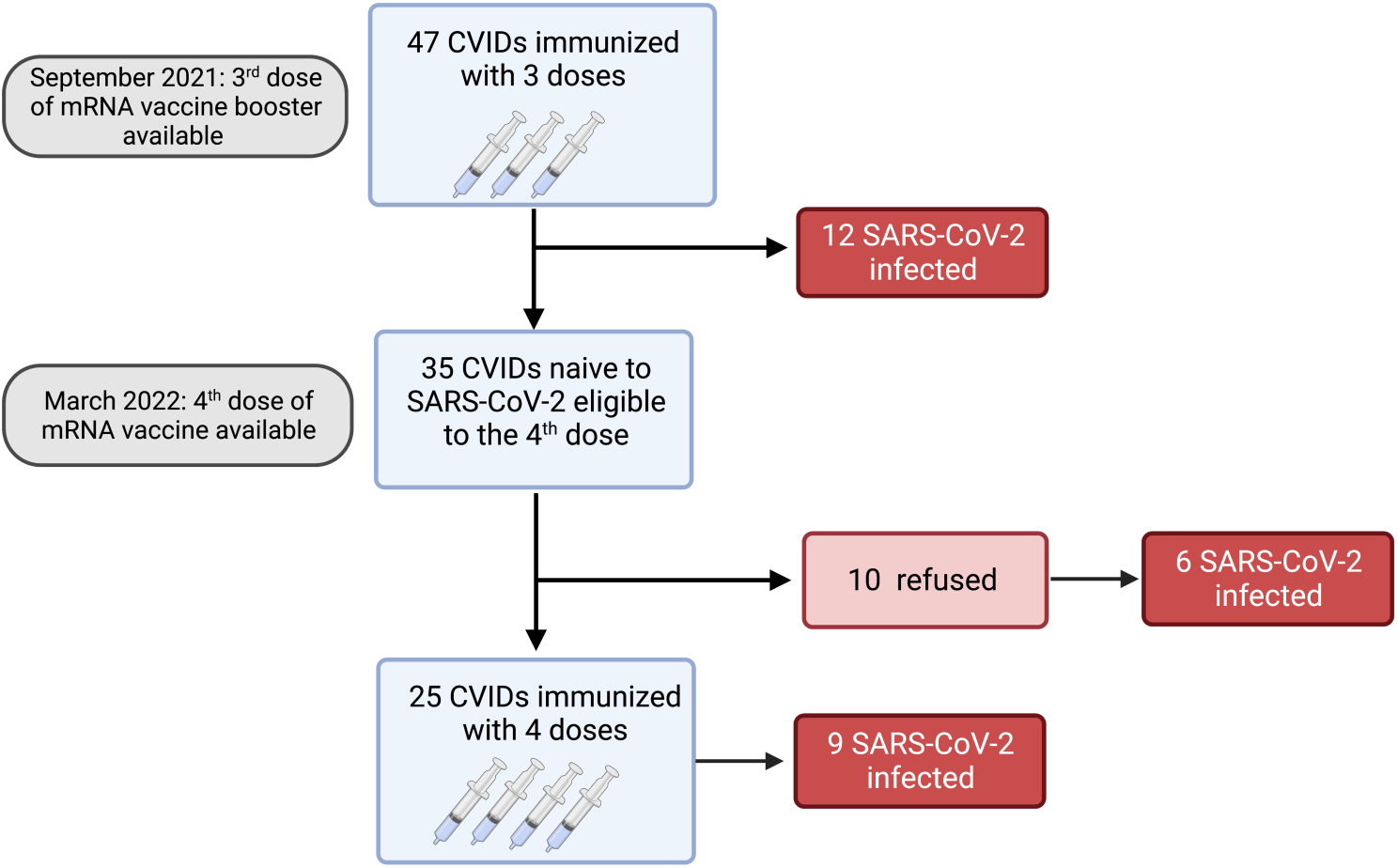
Flow chart. Observational study on 47 adults with CVIDs naïve to SARS-CoV-2 infection immunized with the booster mRNA BNT162b2 vaccine.

### Serum anti-S1 IgG after the first booster vaccine dose

After the 3^rd^ dose, all HCW controls (median age 54.8 years, IQR 43.8-61.9) had measurable levels of serum anti-S1 IgG antibodies. In contrast, specific IgG were above the cut-off value in 30/47 (63.8%) CVIDs patients (median 21.5 OD ratio, IQR 2.7-33.6) (Figure 2A). We separately show the antibody levels in CVIDs patients that exceeded (*Responders, R*) or not (*Non-Responders, NR*) the cut-off value. CVIDs patients able to seroconvert had a highly variable response with a significantly lower antibody concentration than control HCWs (p<0.0001) (Figure 2A). The data suggest that, even when we exclude *Non-Responders* CVIDs patients from the analysis, our cohort includes individuals with different capacities to mount an antibody response.

**Figure 2.**
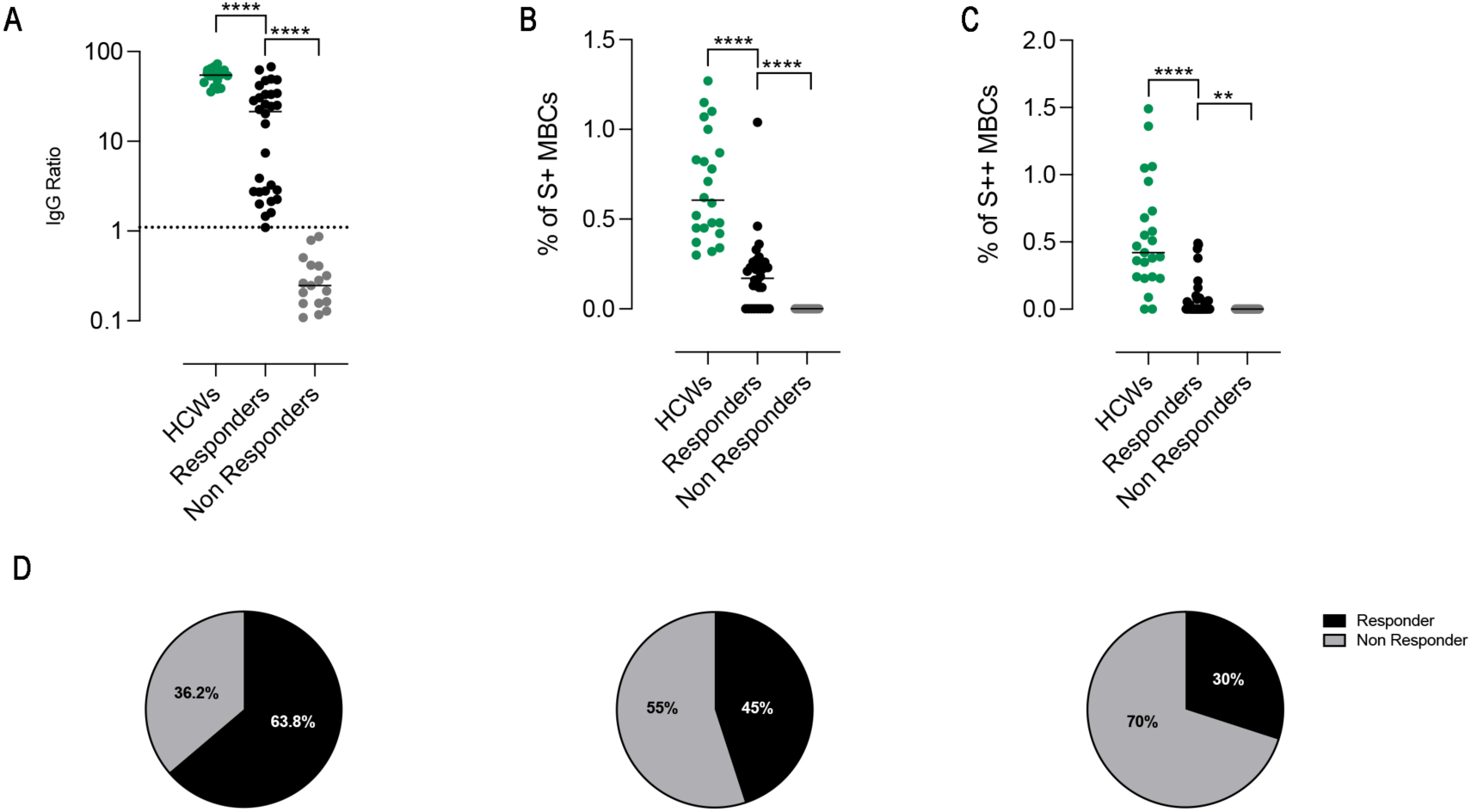
Anti-S1 IgG antibodies and spike-specific memory B cells after the third dose of BNT162b2 vaccine. (A) Individual anti-S1 IgG ratio values in HCWs (green) and in patients with CVIDs. The values measured in the patients with anti-S1 IgG above the cut-off value (dotted line) are shown in black and those below in gray. Bars indicate median values. (B and C) Frequency of low (S+) and high-affinity (S++) MBCs in HCWs (green) and in *Responders* and *Non-Responders* CVIDs patients. (D) The pie chart depicts the percentage of CVIDs patients with serum anti-S1 IgG antibodies above (*Responders*, shown in black) or below (*Non-Responders*, in gray) the cut-off value, measured after the 3^rd^ dose of the mRNA BNT162b2 vaccine and the percentage of CVIDs patients with measurable S+ and S++ MBCs. Non-parametric Mann–Whitney t-test was used to evaluate statistical significance. Two-tailed P value significances are shown; **p< 0.01 and ****p< 0.0001.

### Peripheral blood Spike-specific memory B cells after the first booster vaccine dose

High specificity and affinity are the most important characteristics of MBCs, generated by the adaptive immune system in response to infection or vaccination^25, 37^. MBCs specific for SARS-CoV-2 were identified by their ability to bind the biotin-labeled spike protein. We can distinguish MBCs with low (PE single positive, S+) or high binding capacity (PE-BUV395 double positive, S++) for the spike protein, confirming previously published data^16, 25, 37^ (Supplementary Figure 1A). RBD-specific B-cells were identified by staining with RBD-biotin labeled with streptavidin-FITC (Supplementary Figure 1B). We have shown before that in control HCWs, S+ MBCs can be detected before vaccination, whereas S++ MBCs are produced only after the 2^nd^ vaccine dose^37^. S+ MBCs are mostly of IgM isotype and S++ are switched MBCs (Supplementary Figure 1B). The RBD specificity is absent among S+ MBCs. In contrast, 20-40% of the S++ MBCs bind RBD and are of switched isotype (Supplementary Figure 1B).

We measured S+ (Figure 2B) and S++ MBCs (Figure 2C) ten days after the 3^rd^ dose in patients who were defined as *Responders* or *Non-Responders* based on the level of specific antibodies. Overall, as compared to control HCWs, CVID patients had significantly lower frequencies of S+ and S++ MBCs. S+ MBCs were detectable in 45% (21/47) of the CVIDs patients (Figure 2B) and S++ MBCs in 30% (14/47) of the cases (Figure 2C).

Thus, depending on the biological read-out of efficacy used, a different percentage of patients can be classified as *Responder* or *Non-Responder* (63.8% vs 45% vs 30%) (Figure 2D).

The presence, frequency, and isotype of MBCs have a prognostic value in CVID and identify patients with different severity of the disease^39, 40^. For this reason, we focused our study on MBCs cells generated in response to the COVID-19 vaccine.

Among CVIDs patients who could be considered *Responders* based on antibody levels, we identified three different types of immune responses (Figure 3A and Supplementary Table 2), which were used to define three functional groups of patients. Fourteen patients (group 1) generated both S+ and S++ MBCs, 7 patients (group 2) generated only S+ MBCs and 9 patients (group 3) had no spike-specific MBCs. The 17 *Non-Responder* patients, lacking both antibodies and spike-specific MBCs, were assigned to group 4 (Figure 3A).

**Figure 3.**
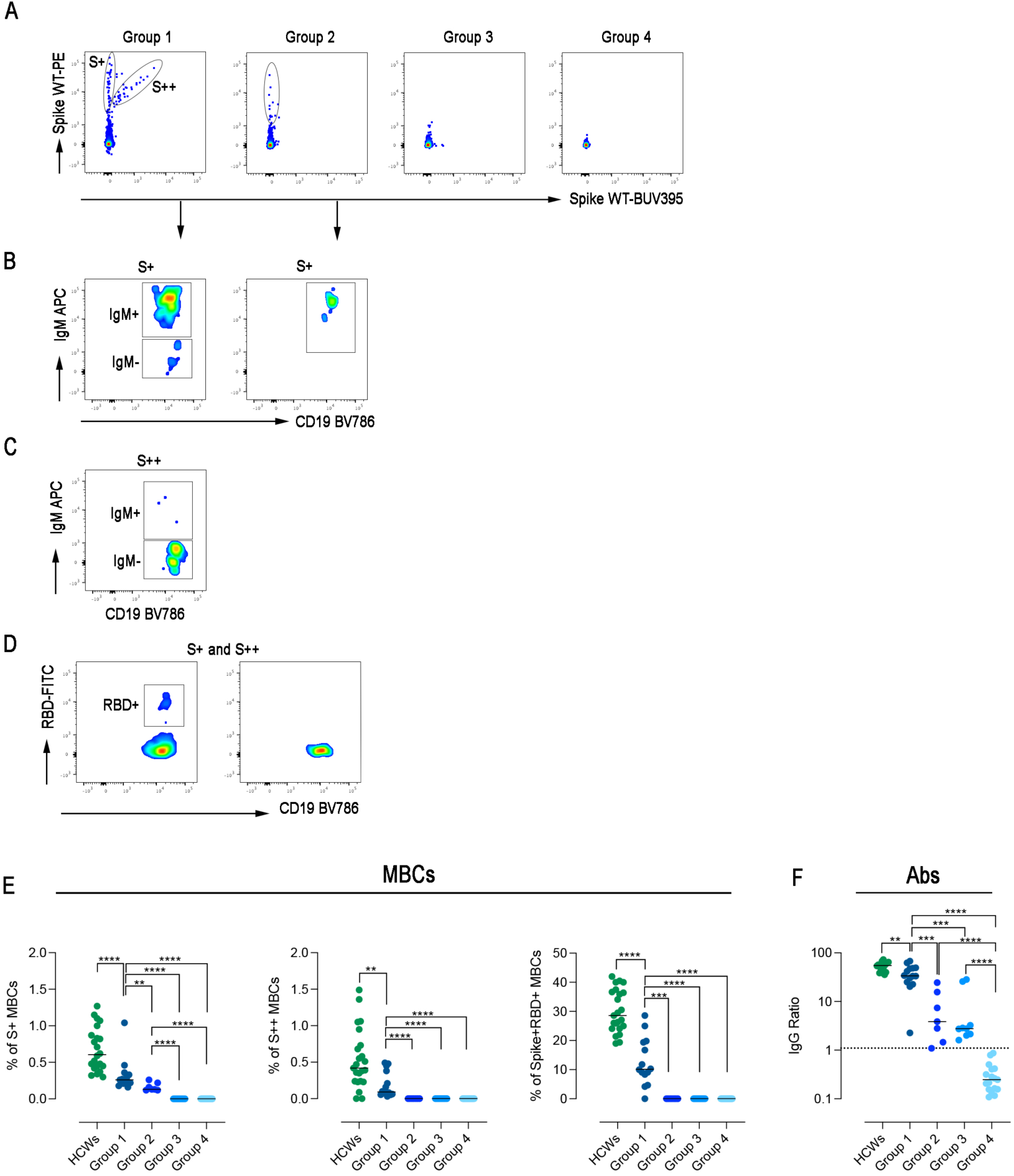
Specific immune response to SARS-CoV-2. (A) Spike-specific MBCs with low (S+) and high (S++) binding capacity are shown in patients representative of the four groups. (B) S+ MBCs, detectable only in patients of groups 1 and 2, are mostly of IgM+ isotype. (C) S++MBCs, found only in group 1, are IgM-MBCs. (D) RBD-specific MBCs were only detectable in group 1. (E) Dot plots represent the frequency of S+, S++, and RBD+ MBCs in the four groups of CVIDs patients and HCWs. (F) The plot shows the concentration of S1-specific IgG in HCWs (in green), and CVIDs patients. Dotted line indicates the cut-off value. Bars indicate median. Non-parametric Mann–Whitney t-test was used to evaluate statistical significance. Two-tailed P value significances are shown as **p< 0.01, ***p< 0.001, ****p< 0.0001.

As described before in control HCWs^37^, S+ MBCs were mostly IgM MBCs and S++ MBCs expressed switched isotypes (Figure 3B and C). Although group 1 patients generated an immune response composed of S+, S++, and RBD+ MBCs (Figure 3D), all frequencies were significantly lower compared to HCWs (p<0.0001; p=0.0037 and p<0.0001) (Figure 3E). In group 2, only S+ IgM MBCs were detectable (Figure 3B), whereas patients of groups 3 and 4 failed to generate any spike-specific MBCs.

Serum anti-S1 IgG levels differed in the four groups, being significantly higher in group 1 than in groups 2 and 3 (p=0.0008 and p=0.0006, respectively) (Figure 3F).

### The immune response after the 4^th^ vaccine dose

Six months after the 3^rd^ dose, the 4^th^ dose was administered to 25 patients. Twelve patients did not receive the second booster because they were infected by SARS-CoV-2 after the 3^rd^ dose (Figure 1) and were not included in the longitudinal study.

After the 4^th^ vaccine dose, in group 1, the frequency of S+, S++ RBD-specific MBCs significantly increased (p=0.0386, p=0.0122, p=0.0245, respectively) (Figure 4A-C). In group 2, the frequency of S+ MBCs did not change, but few S++ and RBD-specific MBCs were generated in four patients (Figure 4B-D). In one patient of group 3, S+ MBCs became detectable after the 4^th^ dose (Figure 4A).

**Figure 4.**
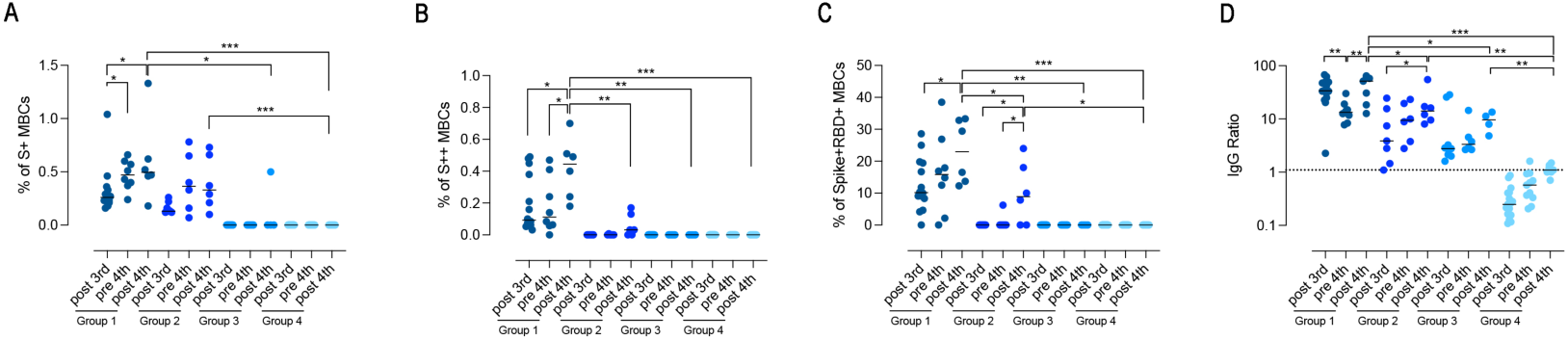
Persistence of the specific response against SARS-CoV-2. (A-D) Graphs show the frequency of spike-specific S+ MBCs (A) S++ MBCs (B), RBD+ MBCs (C) and the concentration of anti-S1 IgG (D), after the 3rd dose, before and 10 days after the 4^th^ dose in the four groups of CVIDs patients. Medians are shown as bars and the dotted line indicates the cut-off. Non-parametric Wilcoxon matched pair signed-rank test and Mann–Whitney t-test were used to evaluate statistical significance. Two-tailed P value significances are shown as * p<0.05, **p< 0.01, ***p< 0.001.

In the pre-4^th^ dose serum sample, anti-S1 IgG levels were still detectable in all patients who had produced antibodies before (groups 1, 2, and 3) (Figure 4D). In group 1, anti-S1 IgG declined 6 months after the 3^rd^ dose (p=0.0012) and increased again in response to the 4^th^ dose (p=0.0093) following the kinetics observed in healthy controls^25^. Anti-S1 IgG increased in response to the booster dose in group 2 (p=0.0312), but not in group 3. In groups 2 and 3, antibody levels were significantly lower than in group 1 before and after the 4^th^ dose (p=0.035 and p=0.0121). The 4^th^ dose had no effect on the level of anti-S1 IgG in patients of group 4 (median 1.095 OD ratio IQR 1-1.35) (Figure 4D).

### Isotype of Spike-specific MBCs

The adaptive immune response to vaccination induces the production of MBCs with variable affinities^41^. Usually, in comparison to those expressing IgM, MBCs of switched isotypes have more somatic mutations, carry the imprint of antigen selection, and have a high affinity for their target antigen^33, 42^.

We compared the isotype of MBCs generated in response to vaccination in HCWs and CVIDs patients of groups 1 and 2. In control HCWs, S+ MBCs were half of IgM and half of switched isotypes, whereas S++ MBCs were almost all switched (Figure 5A and B).

**Figure 5:**
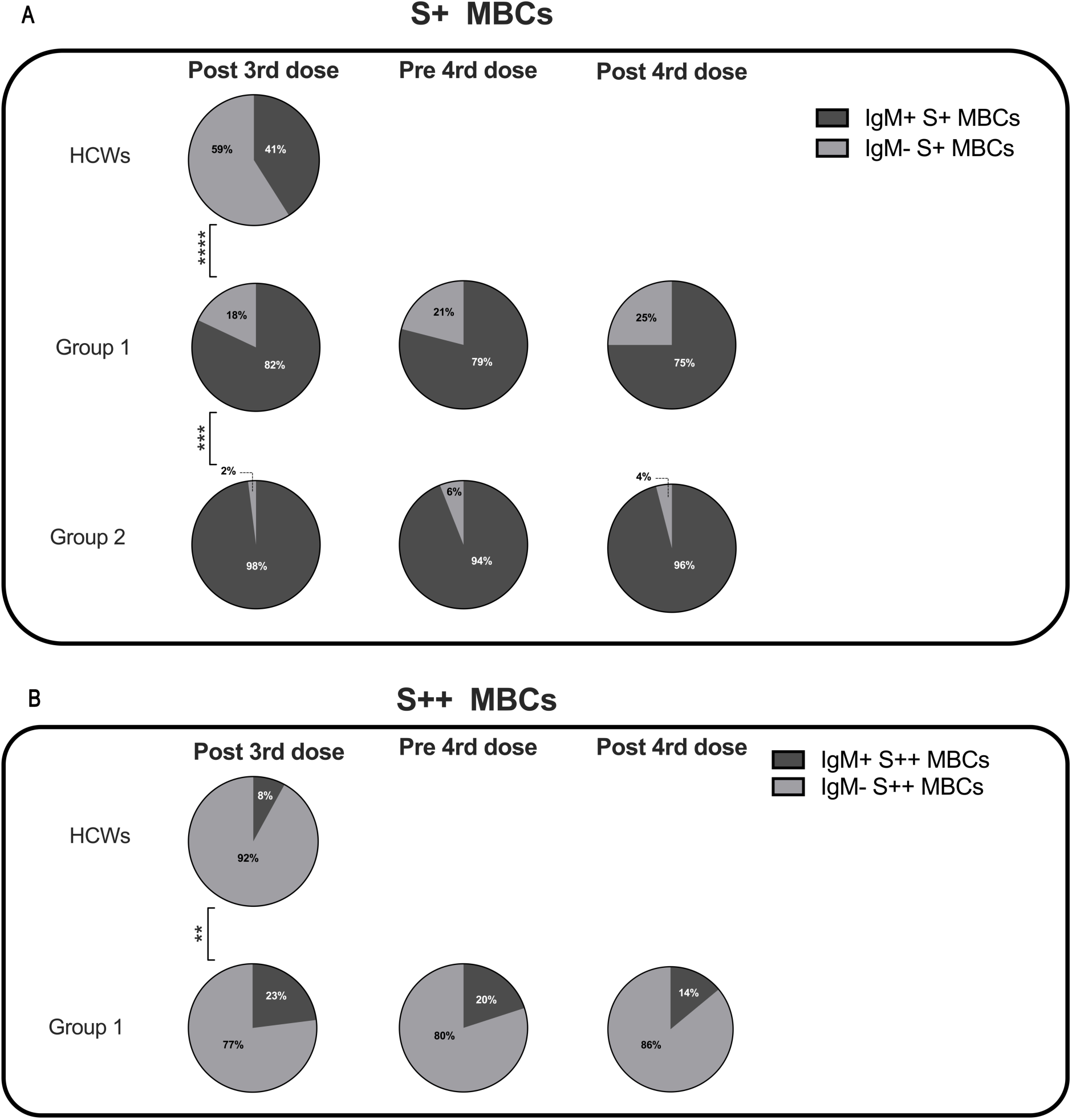
Percentage of IgM+ and IgM-specific MBCs. Pie charts depict the percentage of IgM+ and IgM- S+ (A) and S++ MBCs (B) in HCWs, and groups 1 and 2 CVIDs patients at the time points of analysis. MBCs were always undetectable in group 3 and group 4. Categorical variables were compared by Fisher exact test. Level of significance:**p< 0.01, ***p<0.001, ****p< 0.0001.

Class switching appeared to be less effective in CVIDs patients since both in groups 1 and 2 S+ MBCs were mostly of IgM isotype (Figure 5A). S++ MBCs, detectable only in patients of group 1 at all-time points, were in the majority switched MBCs. The frequency of S++ switched MBCs, however, remained significantly lower than in the controls (Figure 5B).

Thus, in group 1, mRNA vaccination generates all elements of a physiological response: S1- specific serum IgG, S+ and S++ MBCs. However, in comparison to control HCWs, antibody levels are lower, specific MBCs are produced at a reduced frequency and class-switching is impaired. CVIDs patients of group 2, produced even fewer antibodies, and no high-affinity MBCs, but generated S+ and RBD-specific MBCs, all of IgM isotype. No specific MBCs were detectable in groups 3 and 4.

### B-cell phenotype analysis

We asked the question of whether the four groups of CVIDs patients, defined by the response to SARS-CoV-2 vaccination, correspond to distinct B-cell phenotypes. On samples obtained after the 3^rd^ dose, we performed an unbiased analysis of CD19+ B cells using optimized t-stochastic neighbor embedding (Opt-SNE) to reduce dimensionality, followed by graph-based clustering analysis (X-shift). We identified 14 B-cell clusters in the four concatenated groups (Figure 6A). The B cell clusters were analyzed by marker expression to identify the different B cell populations resulting in the definition of seven major clusters (Figure 6B).

**Figure 6:**
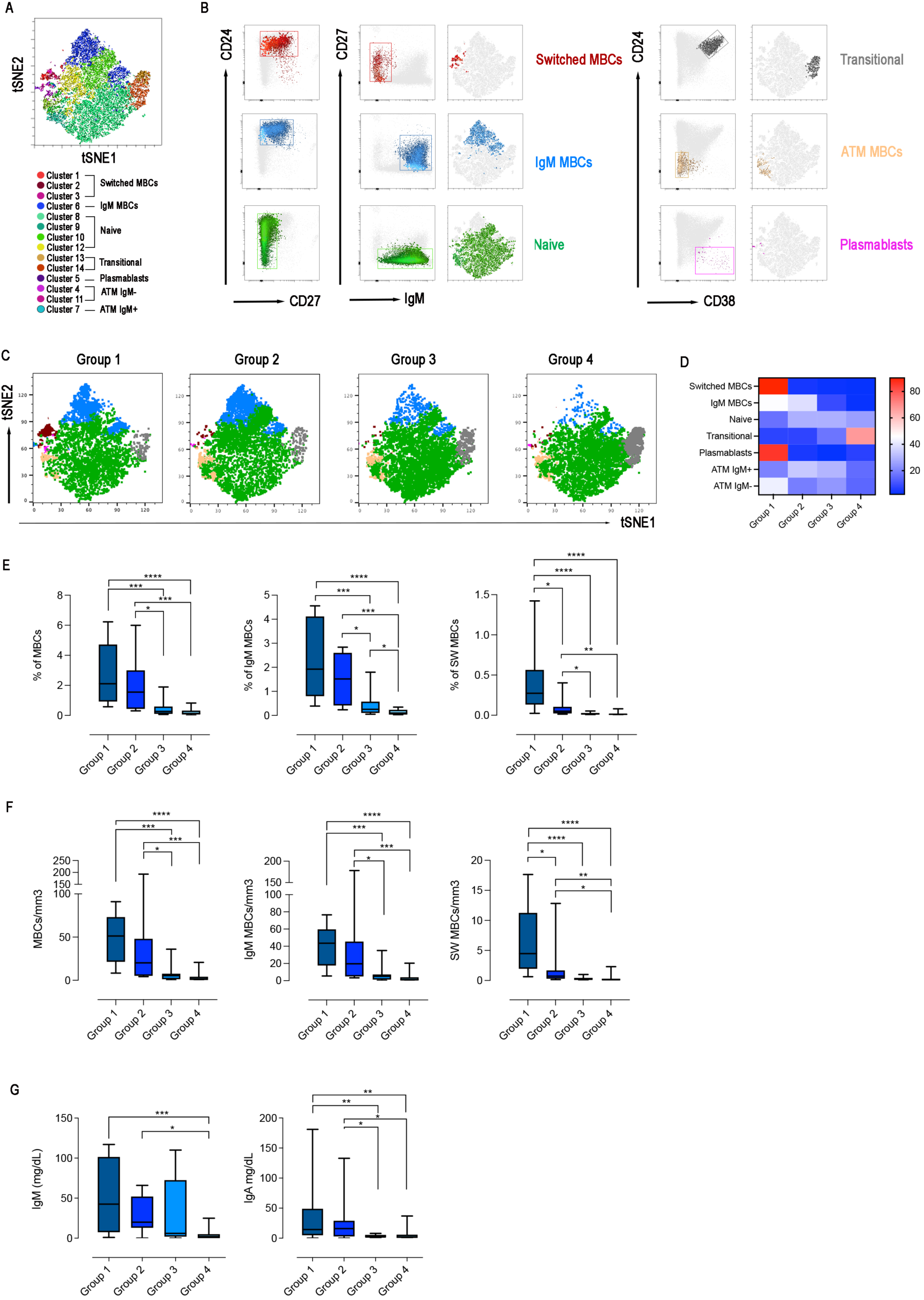
B-cell population in the 4 groups. (A) X-Shift B-cell cluster sets originated from the four concatenated groups and overlaid onto the Opt-SNE map, each cluster is indicated by a color. Based on the relative expression level of surface markers calculated by X-shift, we identified naïve B cells (clusters 11, 12, 13 and 15), switched MBCs (clusters 1, 2 and 3), IgM MBCs (cluster 6), transitional B cells (clusters 13 and 14), plasmablasts (cluster 5), and atypical MBCs IgM- (clusters 4 and 11) and IgM+ (cluster 7). (B) Expression of CD24, CD27, CD38 and IgM for population characterization and identification. For sake of simplicity, we represent the same population as one cluster with one color. (C) Merged Opt-SNE plots for each group with relative X-Shift cluster sets overlaid onto the Opt-SNE map. (D) Heat map depicts the cluster sets abundancy (%) in the four groups. Bar plots depict the frequency (E) and absolute numbers (F) of MBCs (CD19+CD24+CD27+), IgM (IgM+) and switched (IgM-) MBCs in the four CVIDs groups. The frequency of the B cell populations was evaluated in the lymphocyte gate. (G) Serum levels of IgM and IgA in the four groups. Non-parametric Mann–Whitney t-test was used to evaluate statistical significance. Two-tailed P value significances are shown as * p<0.05, **p< 0.01, ***p< 0.001, ****p< 0.0001.

The relative X-Shift cluster sets were calculated for each of the four groups (Figure 6C). The main differences among the groups were caused by the distribution of MBCs.

We found that switched MBCs were present only in group 1, whereas IgM MBCs were identified in groups 1 and 2 (Figure 6C-D). Atypical MBCs were detected at low frequencies in all groups. Plasmablasts were found only in group 1 and transitional B cells were increased in group 4 (Figure 6C-D).

Based on the populations identified by cluster analysis, we calculated the frequency and absolute numbers of the identified populations in all patients of the four groups. Frequency and absolute numbers of MBCs and IgM MBCs were within the normal range in groups 1 and 2 but switched MBCs were significantly reduced in group 2 compared to group 1 (frequency: p=0.0159; absolute numbers: p=0.0297, Figure 6E and F). Patients of group 3 had significantly fewer MBCs, both of IgM and switched isotypes, compared to groups 1 and 2 (Figure 6E and F). Patients of group 4 were lymphopenic and CD19+ B cells were significantly reduced (Supplementary Figure 2A and B). Despite the reduction of total B cells, transitional B cells, the most immature cell type found in the peripheral blood, were present at normal frequency and numbers indicating a normal output from the bone marrow. In contrast, mature-naive B cells and MBCs were strongly reduced, both in frequency and absolute numbers (Supplementary Figure 2A and B, Figure 6E and F). In groups 1, 2 and 3 transitional and mature-naive B cells were within the normal values (Supplementary Figure 2A and B). The frequency and the absolute number of atypical MBCs were comparable in the four groups of patients (Supplementary Figure 2A and B).

In summary, group 1 patients had a B cell phenotype similar to that of control HCWs and all B-cell populations were detected at normal frequencies. Patients of group 2 had normal numbers of IgM MBCs but were characterized by a strong reduction of switched MBCs. Patients in group 3 had no MBCs. Patients of group 4 had low total B-cell numbers with a significant reduction of mature-naive B cells and the absence of MBCs. This distribution remained constant at the three-time points of analysis: after the 3^rd^ dose, before, and after the 4^th^ dose (data not shown).

In line with the description of the B-cell phenotype, IgM serum levels measured at the study time were significantly reduced in group 4 compared with groups 1 and 2 (p=0.0001 and p=0.0238, respectively) and IgA levels were significantly reduced in groups 3 and 4 compared to group 1 (p=0.0052, 1 *vs* 3; p=0.0018, 1 *vs* 4) and group 2 (p=0.0428, 2 *vs* 3; p=0.0500, 2 *vs* 4) (Figure 6C).

### T cell phenotype and function in the four groups of CVIDs patients

The adaptive response to vaccination requires the collaboration of B cells with CD4+ T cells in the GC. We have demonstrated before that switched MBCs can be only generated in individuals with functional GCs^33^. IgM MBCs, instead, have been also detected in individuals with severe immunodeficiencies, lacking T cells or the costimulatory molecule CD40 Ligand (CD40L)^33^. Expression of CD40L by T cells is known to be crucial for affinity maturation and class- switching ^43, 44^. Moreover, the frequencies of specific IFNγ- and TNF*α*-producing CD4+ T cells correlate with the persistence of MBCs specific for the spike protein and with high levels of SARS-CoV-2-neutralizing antibodies^45, 46^.

Because of their importance in the GC reaction and generation of immune memory, we analyzed the function of CD4 T cells activated by polyclonal stimulation in samples obtained after the 3^rd^ dose. We evaluated their phenotype and ability to respond with upregulation of the CD40L and secretion of IFNγ and TNF*α*. The Opt-SNE map identified 7 clusters corresponding to naïve and memory T cells, that expressed or not the CD40L and secreted TNF*α* and/or IFNγ (Figure 7A and B). The relative X-Shift cluster sets were calculated for each of the four groups of CVID patients (Figure 7C). As shown by the heatmap, the function of naïve and memory T cells was maintained in group 1, slightly altered in group 4, but severely impaired in groups 2 and 3 (Figure 7D). Based on the populations identified by cluster analysis, we calculated the frequency and absolute numbers of the identified populations in the 4 groups of patients. T cells of CVIDs group 1 and 2 patients were present in normal frequency and absolute numbers, while total T cells, CD4+ memory (CD45RO+) and CD4+ T cells were low in group 4. One patient of group 3 (11%) and 6 of group 4 (35%) had a severe reduction in CD4+ (<200 cells/mm3) and/or CD4 naive cells (20 cells/mm3), thus fulfilling the criteria for the diagnosis of Late-Onset Combined Immunodeficiency (LOCID)^47^.

**Figure 7:**
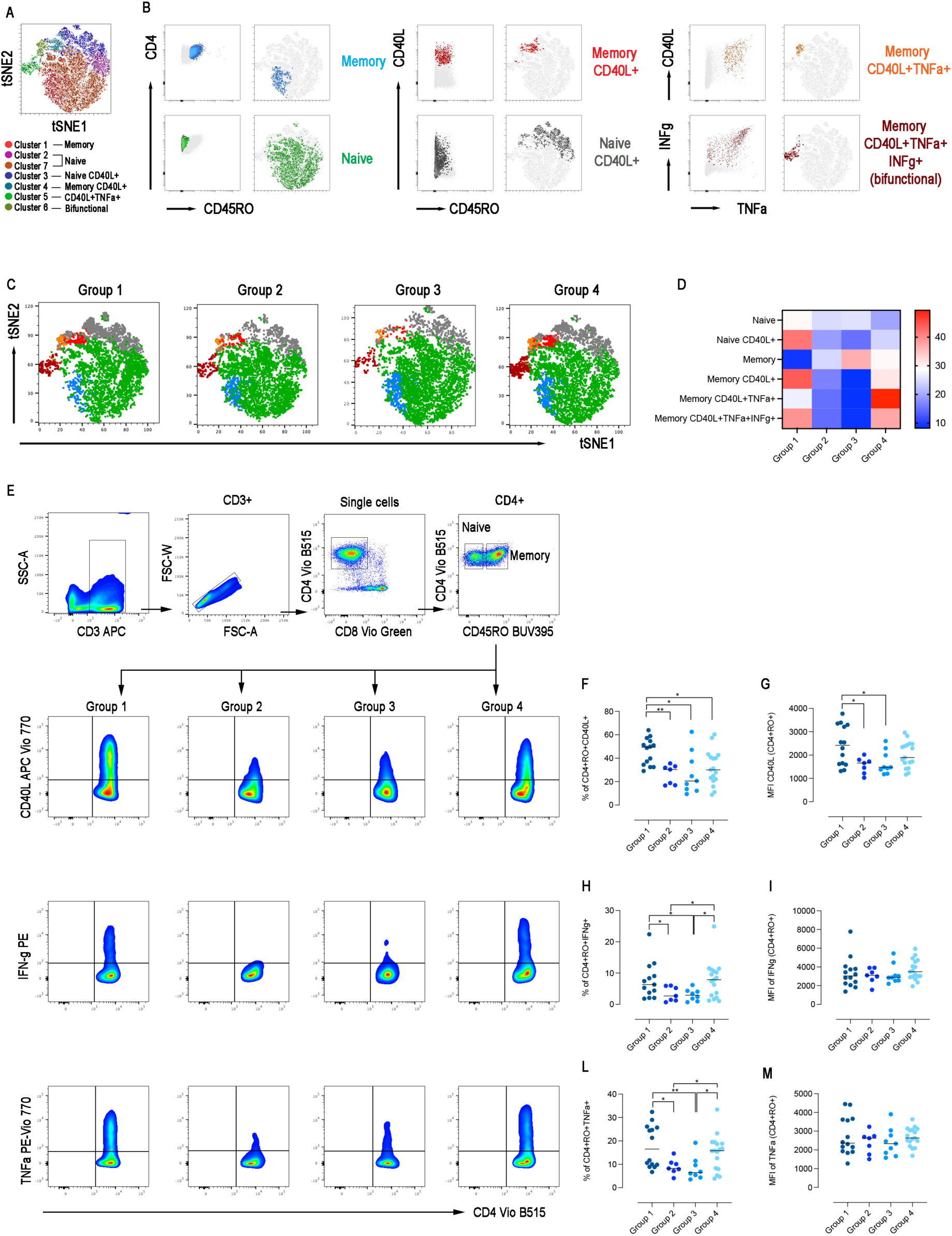
T-cell population in the 4 groups. (A) X-Shift B-cell cluster sets originated from the four concatenated groups and overlaid onto the Opt-SNE map, each cluster is indicated by a color. Naïve T cells were identified in two clusters 2 and 7 and were merged together. (B) Expression of CD45RO, CD40L, IFNg, and TNFa for population characterization and identification. (C) Merged Opt-SNE plots for each group with relative X-Shift cluster sets overlaid onto the Opt-SNE map. (D) Heat map depicts the cluster sets abundancy (%) in the four groups. (E) FACS plots show the gating strategy used to identify CD4+ memory T cells (CD3+CD4+CD45RO+) positive for CD40L, IFNγ and TNFα (intracellular staining) after stimulation with CytoStim in one patient representative of each group. Frequency of CD4+ memory T cells expressing CD40L (F), IFNγ (H) and TNFα (L) in the different groups. Mean Fluorescence Intensity (MFI) of CD40L (G), IFNγ (I) and TNFα (M) (gated on CD4+ memory T cells). All patients were analyzed after the 3^rd^ dose. Bars indicate the median. Non-parametric Mann–Whitney t-test was used to evaluate statistical significance. Two-tailed P value significances are shown as * p<0.05, **p < 0.01.

Upon in vitro polyclonal stimulation, the frequency of CD45RO+ T-cells expressing the CD40L was significantly diminished in groups 2, 3, and 4 compared to group 1 (p=0.0015, p=0.0101, and p=0.0143, respectively) (Figure 7E-F). In groups 2 and 3, also the mean fluorescence intensity (MFI) for the CD40L, which is proportional to the number of expressed molecules, was lower than in group 1 (p=0.0491 and p=0.0327, Figure 7G). T cells of group 4 expressed the same amount of the CD40L of group 1 (Figure 7G). The frequency of CD4+RO+ T cells expressing IFNγ and TNFα after stimulation were significantly reduced in CVIDs patients of groups 2 and 3 compared to both group 1 (IFNγ p=0.0456 and p=0.0266; TNFα p=0.0379 and p=0.0064) and group 4 (IFNγ p=0.0337 and p=0.0266; TNFα p=0.0388 and p=0.0415) (Figure 7E, H and L). The MFI of IFNγ and TNFα was equally high in all groups (Figure 7I and M).

### CVIDs clinical phenotypes

Differences in specific B cell responses between groups are mirrored by the clinical and immunological characteristics of participants (Table 1, and Figure 8A). When compared to groups 3 and 4, groups 1 and 2 had a less severe clinical phenotype, with a significantly lower frequency of patients with signs of immune dysregulation, lower prevalence of chronic lung disease (Figure 8A), and a lower cumulative number of IEI-related disorders (Table 1). None of the patients of groups 1 and 2 showed lymphopenia (<1000 cells/mm3) (Table 1). These observations confirm that the presence of MBCs, not only of switched but also of IgM isotype, is associated with a less severe course of clinical disease^39^. The inability of patients of group 2 to generate high-affinity MBCs, supports the notion that IgM MBCs have the function of systemic and mucosal first-line protection^34, 48^, but do not represent the selected product of adaptive immunity.

**Figure 8:**
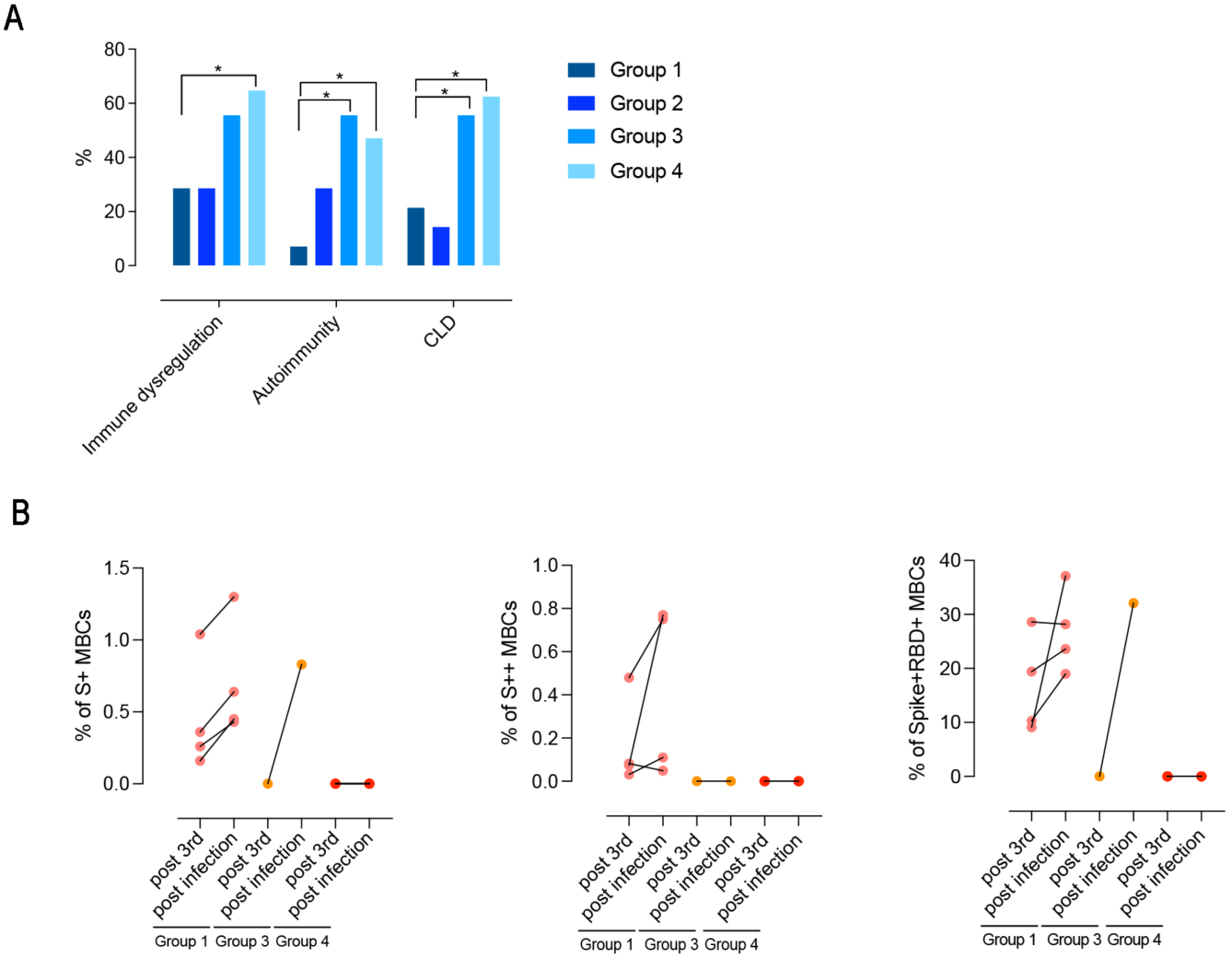
(A) Clinical phenotypes (immune dysregulation, autoimmunity, and CLD) of CVIDs patients grouped according to post immunization S+ and S++ MBCs response and anti-S1 IgG. (B) Paired dot plots depict the S+, S++ and RBD+ MBCs measured before and after an infection in CVID patients who had COVID-19 after 3^rd^ dose. Categorical variables were compared by Fisher exact test. Level of significance: *p<0.05.

**Table 1.**
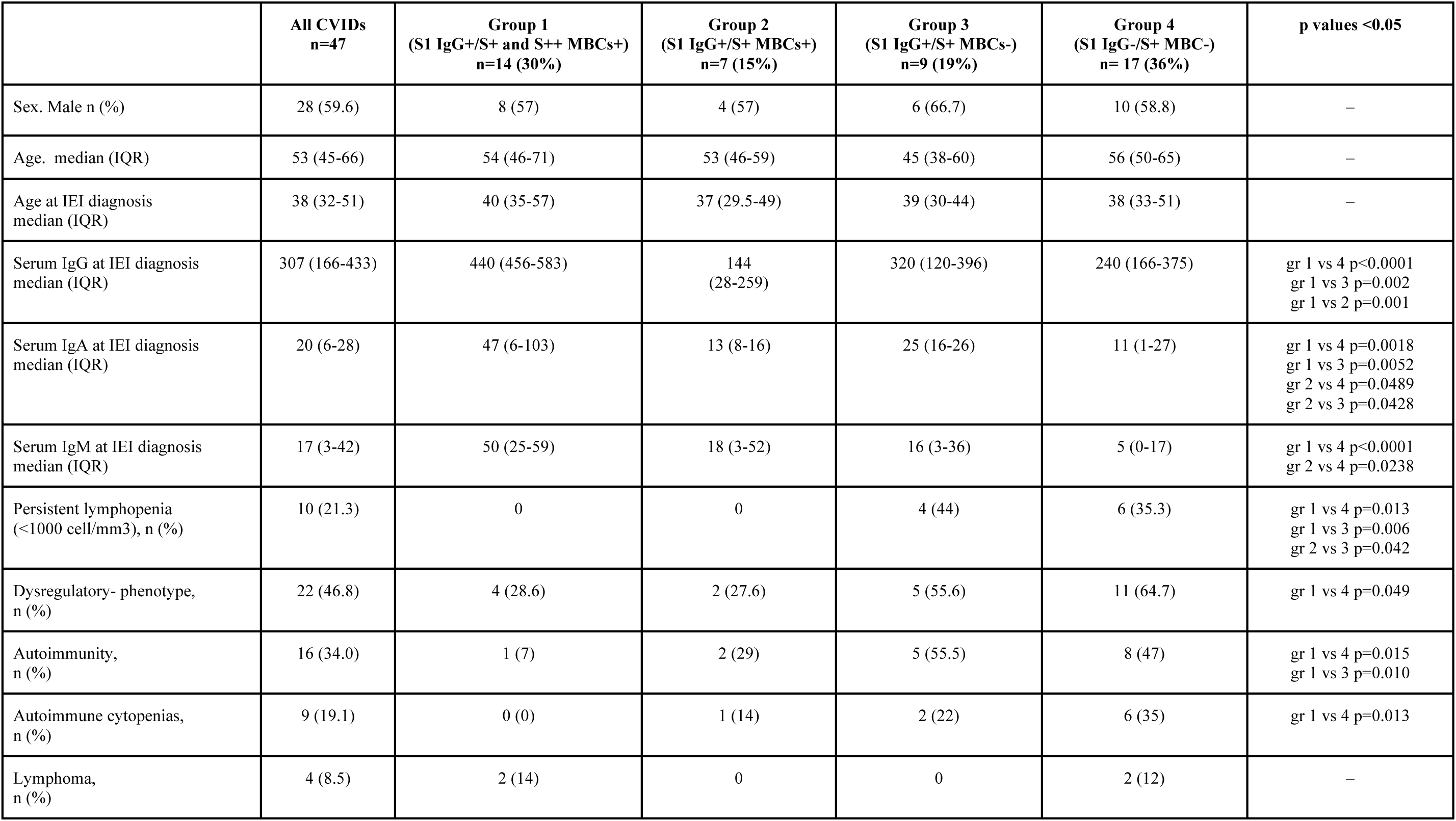
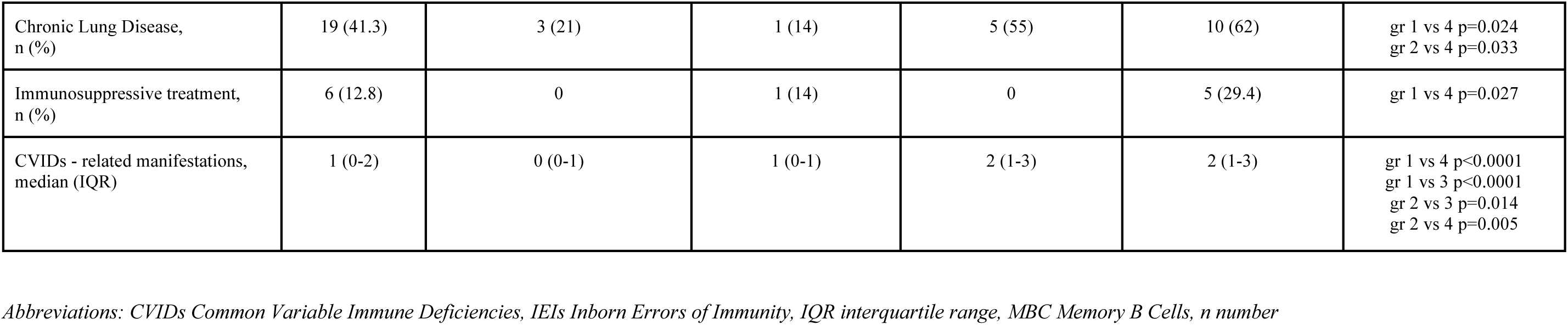
Clinical characteristics and immunological data of 47 CVIDs patients

For 10/47 patients who presented with a severe or complex phenotype, monogenic forms of CVIDs were investigated by targeted gene sequence analysis (Supplementary Table 1). No Pathogenic or Likely Pathogenic Variants were identified, and no molecular diagnosis was achieved. The NGS analysis identified one or more constitutional heterozygous variants classified as Variants of Uncertain Significance (VUS) in four patients (Supplementary Table 3).

### COVID-19 infection post-immunization

After receiving the 3^rd^ dose of the vaccine, 12/47 (25%) patients were infected by SARS-CoV-2. Infected patients were previously classified as being part of group 1, 3 or 4. One month after recovery, the frequency of S+, S++, and RBD+ MBCs increased after infection in group 1. One patient of group 3 generated S+ MBCs of IgM isotype that were able to bind RBD (Figure 8B). Spike-specific MBC analysis revealed that infection reinforces the immune memory generated after the first booster dose in patients of group 1, whereas no response was detectable in patients of group 4.

Nine out of 25 (36%) of CVIDs patients were infected by SARS-CoV-2 after receiving the 4^th^ dose of vaccine, and 6/10 (60%) were infected after refusing the 4^th^ dose (p=0.436).

The vast majority (24/27) of infected patients received a SARS-CoV-2 specific treatment (monoclonal antibodies or antivirals) and none had a severe COVID-19 course. Twenty-six patients (96%) had a mild COVID-19 course, whereas one patient from group 4 who refused to receive the 4^th^ dose died after being infected by SARS-CoV-2 due to the progression of his lymphomas. Details of SARS-CoV-2 infected patients are reported in Table 2.

**Table 2:** COVID-19 infection courses in enrolled patients

| Group | Sex/age<br>(range) | Days of qRT<br>PCR positivity | SARS-CoV-2<br>treatment | Vaccine dose | Infection course |
| --- | --- | --- | --- | --- | --- |
| group 3 | F/80-85 | 26 | MoAbs | 3D | mild |
| group 3 | M/40-45 | 8 | No | 3D | asymptomatic |
| group 4 | F/60-65 | 22 | MoAbs | 3D | mild |
| group 4 | F/60-65 | 41 | MoAbs | 3D | mild |
| group 1 | M/50-55 | 14 | MoAbs | 3D | mild |
| group 4 | F/75-80 | 25 | MoAbs | 3D | mild |
| group 4 | F/70-75 | 16 | MoAbs | 3D | mild |
| group 1 | F/45-50 | 37 | MoAbs | 3D | asymptomatic |
| group 1 | M/55-60 | 26 | MoAbs | 3D | mild |
| group 1 | M/30-35 | 11 | MoAbs | 3D | mild |
| group 4 | M/80-85 | 7 | Antiviral | 3D | mild |
| group 4 | M/60-65 | 8 | MoAbs | 3D | mild |
| group 4 | M/50-55 | 75 | MoAbs | 3D | mild |
| group 4 | M/50-55 | 14 | Antiviral | 3D | asymptomatic |
| group 4 | F/60-65 | 21 | MoAbs | 3D | asymptomatic |
| group 1 | F/40-45 | 26 | Antiviral | 3D | mild |
| group 3 | F/50-55 | 10 | No | 3D | mild |
| group 3 | F/45-50 | 30 | MoAbs | 3D | mild |
| group 3 | F/65-70 | 10 | MoAbs | 4D | mild |
| group 2 | M/60-65 | 17 | MoAbs | 4D | mild |
| group 1 | F/35-40 | 10 | No | 4D | mild |
| group 1 | F/65-70 | 10 | Antiviral | 4D | mild |
| group 4 | M/55-60 | 10 | Antiviral | 4D | mild |
| group 1 | F/70-75 | 9 | Antiviral | 4D | asymptomatic |
| group 1 | M/55-60 | 7 | Antiviral | 4D | mild |
| group 4 | M/50-55 | 10 | Antiviral | 4D | mild |
| group 3 | F/45-50 | 10 | Antiviral | 4D | mild |

## Discussion

The primary immunization cycle against SARS-CoV-2 has proven to be strongly immunogenic and effective in immunocompetent subjects. However, the emergence of viral variants and waning of antibody levels over time prompted consideration for the administration of vaccine booster doses^49^ and the development of variant-adaptive preparations used to boost and broaden the establish immune memory^50^. In vulnerable patients, such as patients with IEIs, immunization with two doses of mRNA vaccines followed by one or two booster doses has become available early in the second year of the pandemic, as recommended by the European Agency for Drugs (EMA) ^51^, due to the underlying higher risk of morbidity and mortality. Published reports showed that after the primary cycle of mRNA immunization, IEIs patients had variable levels of serum antibodies^10^, mainly depending on the associated defect in their immune response. Limited data on specific immune responses are available after the third dose in this population^14, 52, 53^, whereas data on the fourth dose are lacking.

SARS-CoV-2 booster immunizations in humans primarily recruit pre-existing MBCs^54–56^, demonstrating that they are essential for the reaction to antigen re-encounter and responsible for systemic and local protection^25, 57^. In this longitudinal study on adult CVIDs naive to SARS-CoV-2 infection, all of whom were immunized with one or two booster doses of the Pfizer-BioNTech (BNT162b2) vaccine, we combined the analysis of S1-specific IgG and spike-specific MBCs.

We demonstrate that the measurement of specific antibodies is not sufficient to consider a patient *responder* to the vaccine. Moreover, the integration of the data on S1-specific IgG level and spike-specific MBCs frequency identifies different groups of CVIDs patients with immune impairments of increasing severity.

We show that, upon booster vaccination, 63.8% of the patients produce S1-specific IgG, but only 30% generate all expected elements of B cell memory (group 1). Only group 1 patients were able to respond to booster immunizations with a significant increase in specific IgG, S++, and RBD-specific MBCs.

Fifteen percent of patients with CVIDs (group 2) produced low levels of S1-IgG and generated S+ MBC of the IgM isotype that did not evolve into switched MBC, their more sophisticated successors^33, 42^. Serum S1-specific IgG increased slightly, but MBCs did not change in response to the 4^th^ dose.

More than half of the CVIDs participants were unable to generate specific MBC cells after booster doses: 19% of the patients (group 3), although they generated low levels of anti S1- IgG, completely lack all MBC, while 36% of the patients (group 4), did not develop either S1- IgG or specific MBC.

Based on the performance in response to vaccination, B cell phenotype, and T cell function, we conclude that CVIDs patients of group 1 are able to generate a functional immune memory. Further studies are necessary to establish whether the antibody deficiency in these patients may be explained by a defect in the generation or persistence of long-lived memory plasma cells, the population responsible for the maintenance of high-affinity and durable antibody responses (Figure 9). Group 2 patients produce low levels of specific IgG and low-affinity MBCs of IgM isotype that, as shown before, are generated by a T- and GC-independent mechanism^32, 33^. In group 2, the impaired capacity of T cells to express the CD40L suggests that the GC reaction may be defective, thus explaining the absence of high-affinity MBCs and the impairment of class-switching. (Figure 9). Group 3 patients may have a combined defect, with B cells unable to undergo T-independent development into IgM MBCs and T cells do not express the CD40L and the cytokines IFN*γ* and TNF*α*. Finally, in group 4, a B cell-intrinsic defect may alter survival beyond the transitional stage of B cells and abolish differentiation in more mature populations, while T cell function is preserved (Figure 9). Several monogenic and polygenic defects have been identified in CVIDs. Future studies will demonstrate whether the four functional groups of patients identified here are characterized by different disease-causing mutations affecting the mechanisms that we hypothesize.

**Figure 9:**
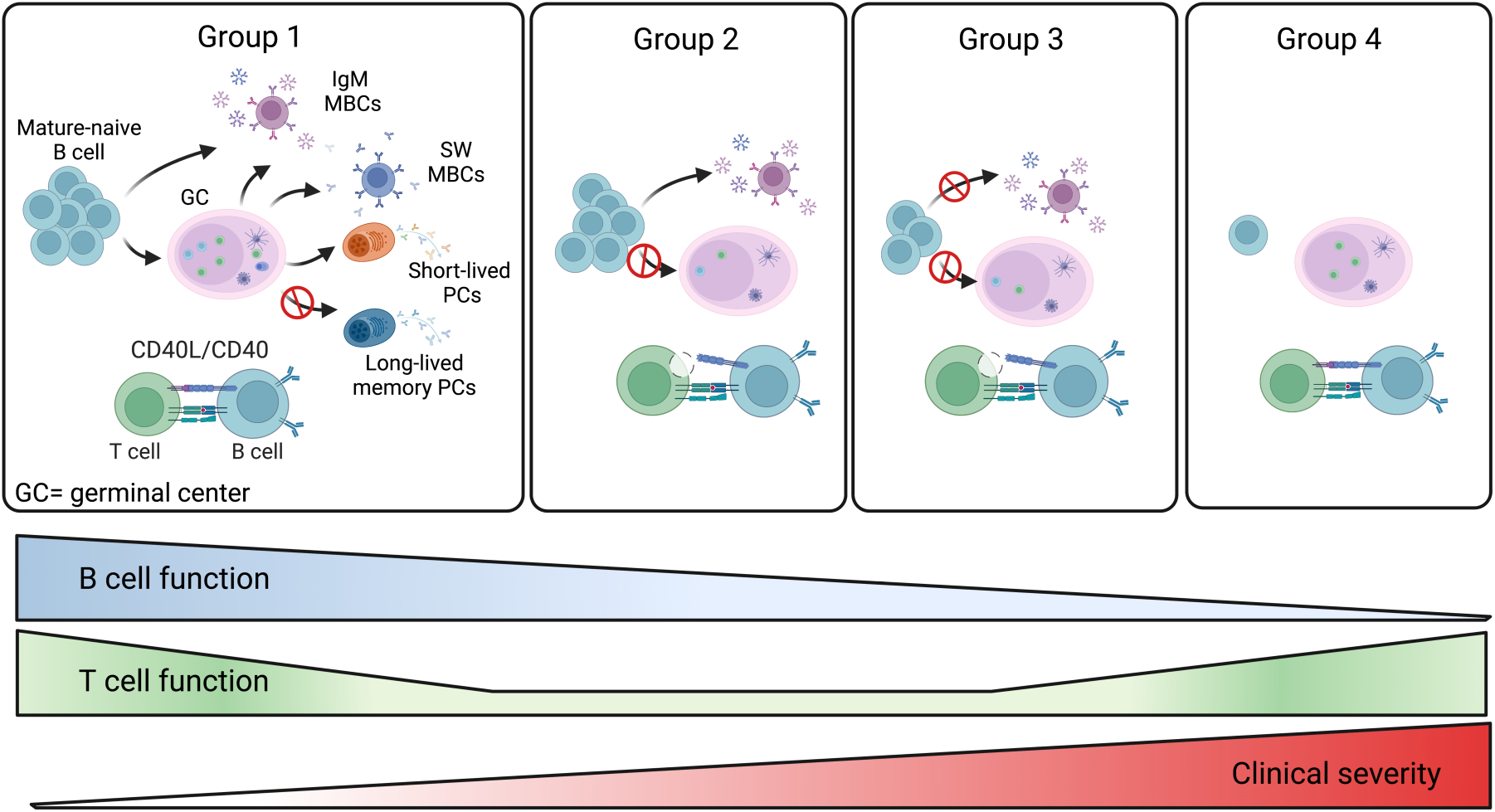
Summary of the four CVIDs groups. The memory B-cell compartment includes GC- and T-independent IgM MBCs, as well as post-GC and T-dependent MBCs of both IgM and switched isotypes. Short-lived plasmablasts (PB) are generated by recent immune responses. Long-lived plasma cells (LLPCs) maintain serum antibody levels in time. Group 1 patients are able to produce all MBCs types. Hypogammaglobulinemia may be explained by the inability to produce LLPCs in this group of CVIDs patients. For patients of groups 2 and 3, the defective T-cell function may impair GC development. Patients of group 2 retain the ability to generate T-independent IgM MBCs, whereas all MBCs are missing in group 3. In patients of group 4, the significant reduction of all B-cell populations past the transitional B-cell stage suggests a severe defect in all B-cell functions. The severity of clinical disease increases in groups 3 and 4.

The immunological heterogeneity of the immune defects associated with CVIDs has been widely studied^58–60^. It has been shown before that CVIDs patients with normal numbers of IgM and switched MBCs have less severe disease. The frequency and function of peripheral IgM MBCs are predictive of the T-independent response to polysaccharide vaccines and clinical outcomes^6, 39^ and, as confirmed here, in the absence of IgM MBCs, patients with CVIDs have a more severe disease with a significantly higher frequency of chronic lung disease and immune dysregulation^39^. Moreover, IgM MBCs may play a crucial role in COVID-19, as monoclonal antibodies derived from IgM MBCs from patient’s convalescent from the infection with the Wuhan virus are potent neutralizers not only for the wild-type strain, but also for its emerging viral variants^38^. Similarly, antibodies derived from IgM MBCs were able to neutralize the H1N1 influenza virus to which the donor had never been exposed^61^. Here we confirm, however, that specific MBCs, able of recall responses after a renewed encounter with the antigen, is established only if switched MBCs can be generated. Thus, the first-line of protection ensured by IgM MBCs is sufficient to protect patients from severe CVIDs disease in the absence of highly specific B–cell immunity. The IgRT may be sufficient to replace the function of switched MBCs in patients whose MBCs are all of IgM isotype.

When we evaluated the incidence of breakthrough infection, we found that although the number of recorded episodes was too low to reach statistical significance, the rate of SARS-CoV-2 infection in CVIDs who received the second booster was half of those immunized with a single booster dose.

To avoid bias of inclusion, for those with complex/atypical phenotypes, monogenic forms of CVIDs and phenocopies were excluded by NGS analysis for a panel of genes involved in monogenic disease with defects in humoral immunity. However, the possibility of structural variants and non-coding pathogenic variants in the analyzed genes, or the presence of causative variants in genes not featured in the targeted panel cannot be excluded.

In summary, our data indicate that in CVIDs patients, the presence of antibodies alone may not be sufficient to demonstrate the establishment of immune memory after vaccination. Thus, a more complex biological read-out of vaccine efficacy comprising specific antibodies, and B and T cell memory responses may be necessary to measure the immune status of these vulnerable patients.

Several critical insights relevant to the future immunization strategies for CVIDs patients can be drawn from this study. Patients with a residual capacity to respond to vaccines require primary cycles of immunization, followed by booster doses due to the critical contribution of repeated antigen exposures to strengthen their responses. On the contrary, in *Non-responders* additional booster doses are unable to induce B-cell memory. Specific T cell responses are also defective in most cases^14^. Thus, replacement by specific monoclonal antibodies as therapy and prophylaxis, and antiviral agents is necessary to limit the viral spread, morbidity, and mortality^62^.

Similarly to patients with CVIDs, defective immune responses to COVID-19 vaccines have been demonstrated in patients under immune suppressive therapy^63^, undergoing treatments for cancer ^64^, and after hematopietic stem cell transplantation^65^. Our results indicate that the detection of specific MBC responses combined with B and T cell phenotypes is a more adequate indicator of vaccine immune efficacy than antibody measurement in vulnerable patients. This analysis may not only identify individuals that remain unprotected after vaccination but also address the search for the underlying immune defect.

## Data Availability

Resource Availability.
Lead Contact
Further information and requests for resources and reagents should be directed to and will be fulfilled by the lead contact, Rita Carsetti
Materials Availability
All reagents will be made available on request after completion of a Materials Transfer Agreement.

## Acknowledgment

The study was supported by funding of the Italian Ministry of Health COVID2020-12371817 and by Grant ‘‘5 per mille, 2021’’.

## Author contributions

EPM, VM, CA, ST, AFS, SDC, MG, SM, and FS acquired the data. EPM, FP, IQ, and RC were responsible for the analysis of the data and drafted the article. SDC, EPM, and RC performed the unsupervised analysis. AC, CQ, FL GDN, DG, ES, CM, GG, AMP, LB, and SZ critically revised the manuscript for important intellectual content. All authors approved the final version to be published and agree to be held accountable for all aspects of the work. RC, and FL made substantial contributions to the acquisition of funding.

## STAR methods

### Resource Availability

#### Lead Contact

Further information and requests for resources and reagents should be directed to and will be fulfilled by the lead contact, Rita Carsetti

## Materials Availability

All reagents will be made available on request after completion of a Materials Transfer Agreement.

### Study design and patients

This longitudinal study was carried out on 47 adults with CVIDs diagnosed according to the ESID criteria^66^ and regularly followed by the Care Center for adults with IEIs in Rome, Italy. As healthy controls, 22 Health Care Workers (HCWs) were enrolled at the Bambino Gesù Children’s Hospital.

HCWs and patients were considered eligible for the study if they were naïve to SARS-CoV-2 infection at the time of enrollment, had completed the primary schedule with the mRNA BNT162b2 vaccine, and agreed to receive the 3^rd^ dose.

In CVIDs, the first booster (3^rd^ dose) dose was performed in March 2022, six months after completing the full primary immunization schedule. In HCWs, the first booster dose was performed nine months after the last dose of the vaccine.

As prescribed for patients with IEIs by the Italian Agency of Drugs (AIFA), the second booster dose (4^th^ dose) of the BNT162b2 vaccine was offered in September 2022 only to those participants who remained free from SARS-CoV-2 infection since the last vaccine administration. Patients were tested for SARS-CoV-2 infection by RT-PCR on the nasopharyngeal swab (NPS) every time they attended a hospital site, in case of positive household contacts irrespective of symptoms, and upon onset of symptoms possibly related to COVID-19. The duration of the viral shedding was evaluated by recording the dates of the first positive and first negative NPS. In patients with SARS-CoV-2 infection, we recorded COVID- 19 severity (scored according to WHO stage^67^), hospitalization, vaccination status, and SARS-CoV-2 treatments).

Blood samples were obtained 10 days after the 3^rd^ dose (post 3^rd^), six months apart on the day of the 4^th^ dose (pre 4^th^), and 10 days after the 4^th^ dose (post 4^th^). For those who were infected after the 3^rd^ dose, blood samples were collected one month after recovery and separately analyzed. For those who refused the 4^th^ dose of vaccine, blood samples were collected six months after the booster immunization and then included in the analysis as pre 4^th^ values (Figure 1). During the study, participants were allowed to continue their treatments, including IgRT as standard therapy for the underlying antibody deficiency. At the study time, all IgRT brands in use did not contain anti-S1 IgG antibodies^68, 69^. For each participant, clinical data were collected, including age, immunoglobulin levels at IEIs diagnosis, and the number of IEI-related disorders. We defined CVIDs with immune-dysregulation phenotype as having at least one of the following: systemic autoimmunity or autoimmune cytopenia, enteropathy, lymphoproliferative disorders, and lymphoid malignancy or solid cancer. Chronic Lung Disease (CLD) was defined as having at least one of the following: Chronic Obstructive Pulmonary Disease diagnosed according to GOLD guidelines^70^, Interstitial Lung Disease or Bronchiectasis (based on CT scan imaging), and Granulomatous and Lymphocytic Interstitial Lung Diseases (GLILD) (based on CT scan imaging and histologically confirmed)^71^. Immunological laboratory data collected at baseline included complete blood count (CBC) and immunoglobulin serum levels.

Following the actual position of IUIS^1^, ten patients displaying complex clinical phenotypes (mainly immune dysregulation) underwent molecular testing to rule out monogenic forms of humoral immunodeficiencies. A custom targeted Next Generation Sequencing (NGS) panel of 53 genes whose variants are associated with CVIDs, and humoral immunodeficiency was performed (Supplementary Table 1). The study was approved by the Ethical Committee of the Sapienza University of Rome (Prot. 0521/2020, July 13, 2020) and performed in accordance with the Good Clinical Practice guidelines, the International Conference on Harmonization guidelines, and the most recent version of the Declaration of Helsinki.

### Anti-S1 IgG antibodies

A semi-quantitative in vitro determination of human IgG antibodies against the SARS-CoV-2 (IgG-S1) was performed on serum samples by using the Anti-SARS-CoV-2 Spike ELISA (EUROIMMUN), according to the manufacturer’s instructions. Values were then normalized for comparison with a calibrator. Results were evaluated by calculating the ratio between the extinction of samples and the extinction of the calibrator. Results are reported as the ratio between the OD of the sample and the OD of the calibrator. The ratio interpretation was as follows: <0.8 = negative, ≥0.8 to <1.1 = borderline, ≥1.1 = positive.

### Cell Isolation and Cryopreservation

Peripheral blood mononuclear cells (PBMCs) were isolated by Ficoll Paque™ Plus 206 (Amersham PharmaciaBiotech) density-gradient centrifugation and immediately frozen and stored in liquid nitrogen until use. The freezing medium contained 90% Fetal Bovine Serum (FBS) and 10% DMSO.

### Detection of antigen-specific B cells

For the detection of SARS-CoV-2 specific B cells, biotinylated protein spike was individually multimerized with fluorescently labeled streptavidin as previously described^16, 25, 37^. Separate aliquots of recombinant biotinylated Spike or Receptor Binding Domain (RBD) were mixed with streptavidin BUV395, streptavidin PE or streptavidin FITC (BD Bioscience) at 25:1 ratio 20:1 ratio and 2.5:1 ratio respectively. Streptavidin PE-Cy7 (BD Bioscience) was used as a decoy probe to gate out SARS-CoV-2 antigen non-specific streptavidin-binding B cells. The antigen probes individually prepared as above were then mixed in Brilliant Buffer (BD Bioscience). 4x10^6^ previously frozen PBMC samples were prepared and stained with the antigen probe cocktail containing 100 ng of spike per probe (total 200 ng), 27.5 ng of RBD, and 2 ng of streptavidin PE-Cy7. After one step of washing, surface staining with antibodies was performed in Brilliant Buffer at 4 C° for 30 min. B cell subsets were identified based on the expression of CD19, CD27, CD24, and CD38 markers by flow-cytometry. Naïve B cells were identified as CD19+CD24+CD27-, transitional B cells were gated as CD19+CD24++CD38++, and atypical MBCs as CD19+CD24-CD27- CD38-. MBCs were defined as CD19+CD24+CD27+, IgM, and switched MBCs were discriminated based on the expression of the IgM. Spike-specific MBCs were defined as low-affinity (positive for PE, S+) or high-affinity (double positive for PE and BUV395, S++) (Supplementary Figure 1A).

Among S+ and S++ MBCs, IgM and RBD expressions were evaluated (Supplementary Figure 1B).

Stained PBMC samples were acquired on FACs LSRFortessa (BD Bioscience). At least 2x10^6^ cells were acquired and analyzed using Flow-Jo10.8.1 (BD Bioscience). Phenotype analysis of antigen-specific B cells was only performed in subjects with at least 10 cells detected in the respective antigen-specific gate. Blank was determined in unexposed donors, before vaccination. LOB (limit of blank) was set as the mean of the blank + 1.645xSD. LOD (limit of detection) as the mean of the blank + 2xSD.

### T-cell functional studies

PBMCs isolated from CVIDs patients were cultured in 96-well cell plates at 1x10^6^ cells/well concentration in RPMI 1640 culture medium containing 5% AB serum. PBMCs were cultured at 37 C°, 5% CO_2_, with CytoStim® (from the SARS-CoV-2 Prot_S PBMC Kit, Miltenyi, Biotech) for a total of six hours. After two hours of incubation, Brefeldin A (SARS-CoV-2 Prot_S PBMC Kit, Miltenyi, Biotech) was added to the cells to inhibit the transport of proteins to the cellular membrane. Cells were fixed, permeabilized, and stained with CD3, CD4, CD8, CD154 (CD40L), IFNγ, and TNFα (SARS-CoV-2 Prot_S PBMC Kit protocol, Miltenyi Biotech) with the addition of CD45RO (BD Biosciences).

T cells were gated as CD3+ and divided into CD3+CD4+ T cells. Naïve T cells were gated as CD3+CD4+CD45RO- and memory T cells were identified as CD3+CD4+CD45RO+. The CD40L, IFNγ, and TNFα were evaluated on CD4 memory T cells. Cells were acquired on a BD FACSymphony A5™ and data were analyzed with FlowJo ver. 10.8.1 (BD Bioscience).

### B cells and T cells unbiased population identification

FCS files were analyzed using the FlowJo ™ v10.8.1 software (BD, Biosciences). Prior to analysis, all samples were normalized to equal cell numbers. For each of the four groups of patients, 5 randomly selected samples were concatenated in a single FCS file. Equal numbers of live CD19+ B cells or CD3+CD4+ T cells were analyzed. Data were partitioned into clusters in high dimensional space by using the X-Shift plugin algorithm^72^, and visualized after performing dimensionality reduction using the Optimized t-distributed stochastic neighbor embedding (opt-SNE). Marker expression and frequency across different clusters were then assessed using the heatmaps generated by the Cluster explorer plugin (FlowJo ™ v10.8.1 software, BD, Biosciences).

### Genetic analysis

Sequencing experiments were performed on the Ion Torrent S5 platform (Thermo Fisher Scientific). Sequence data analysis was performed with the Ion Reporter v5.12 software (Thermo Fisher Scientific). Variant validation was performed by Sanger sequencing on the 3730 Genetic Analyzer platform (Thermo Fisher Scientific). Variants with a minor allele frequency >0.01 were filtered out. Only nonsynonymous variants and splice site variants were retained. A visual analysis of the reads including candidate variants mapped against the reference genome (version GRCh37/hg19) was performed on the Integrative Genome Viewer software (IGV, https://software.broadinstitute.org/software/igv/). Variants were classified according to the American College Of Medical Genetics And Genomics (ACMG) guidelines^73^.

### Statistical Analysis

Data were analyzed by GraphPad Prism 9 version 9.3.1 and Statistical Package for Social Sciences version 15 (SPSS Inc). The data were first tested for normality and homoscedasticity using Shapiro-Wilk and Levene’s tests and since the assumptions were violated, non-parametric tests were used for the analysis.

Demographics were summarized with descriptive statistics (median and IQR for continuous values). Immunological and clinical variables were compared between the different study times. A univariate analysis assessed the impact of variables of interest. Values were compared by the non-parametric Kruskal–Wallis test and, if not significant, the Wilcoxon matched pair signed-rank test or the two-tailed Mann–Whitney U-test were used. Categorical variables were compared by Chi-square test. Differences were deemed significant when *P* < 0.05).

---

**Supplementary Figure 1.**
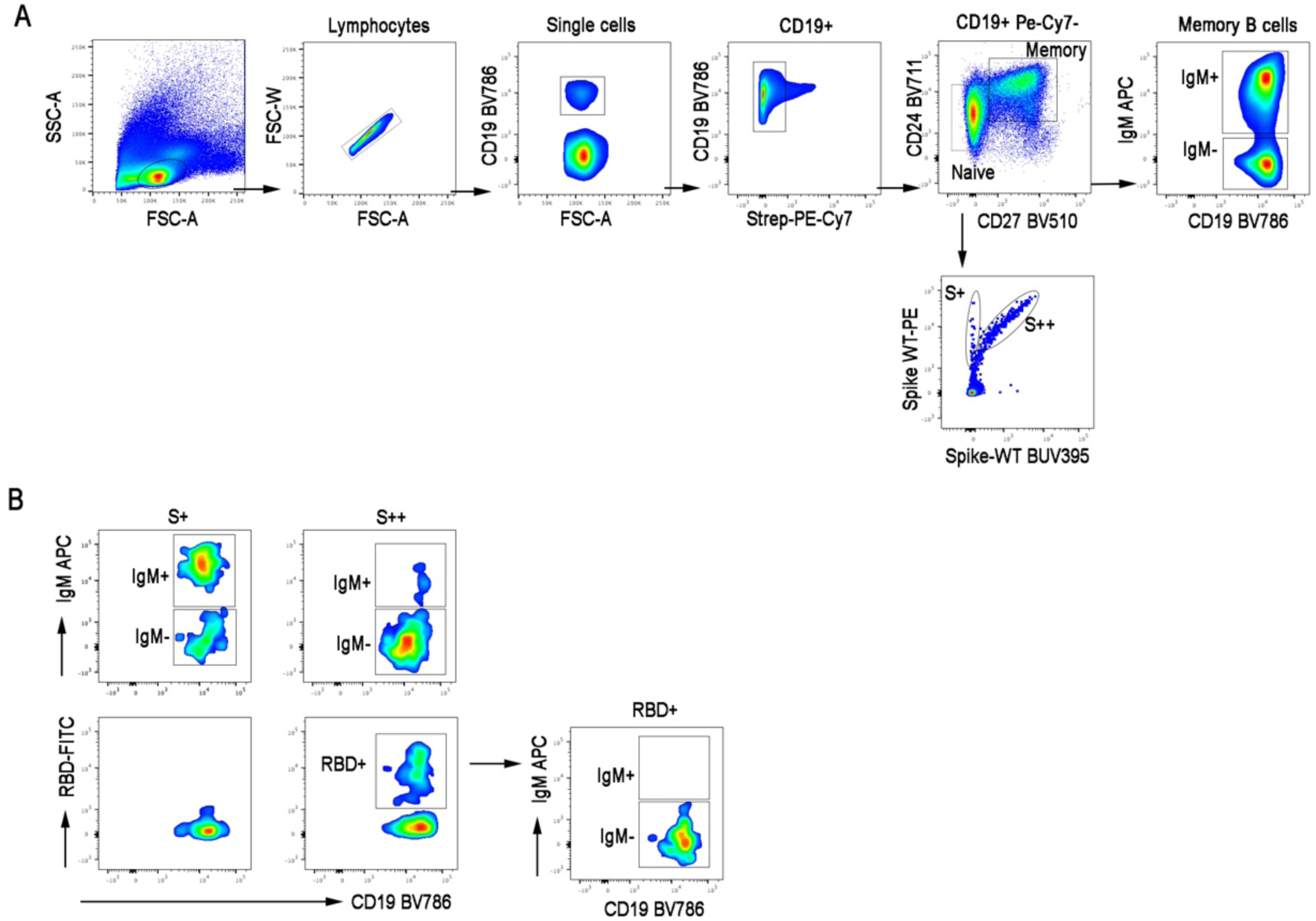
Gating strategy. (A) FACS plots depict the gating strategy for the identification of total (CD19+CD24+CD27+), IgM+ and IgM- MBCs, in a representative HCW. Low (S+) and high (S++) affinity spike-specific MBCs are shown. (B) IgM+, IgM- and RBD+ MBCs among S+ and S++. IgM expression on RBD+ MBCs is shown.

**Supplementary Figure 2.**
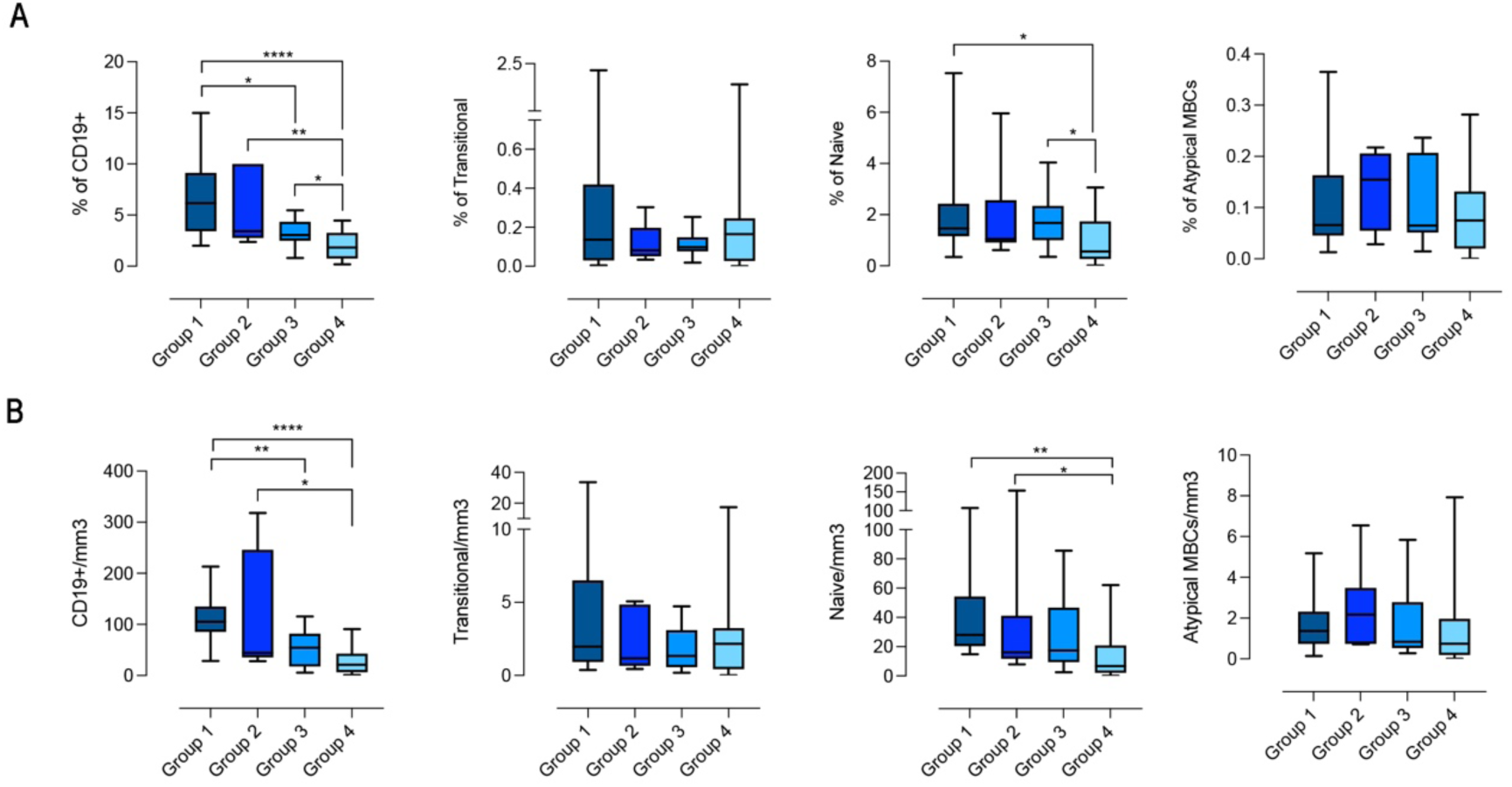
Frequency and absolute B-cell number in the 4 groups. (A-B) Bar plots depict the frequency (A) and the absolute numbers (B) of total B cells (CD19+), transitional B cells (CD19+CD24++CD38++), mature-naive B cells (CD19+CD24+CD27-) and atypical MBCs (CD19+CD24-CD27-CD38-). The frequency of the B cell populations was evaluated in the lymphocyte gate. Non-parametric Mann–Whitney t-test was used to evaluate statistical significance. Two-tailed P value significances are shown as * p<0.05, **p < 0.01, ****p < 0.0001.

**Supplementary Figure 3.**
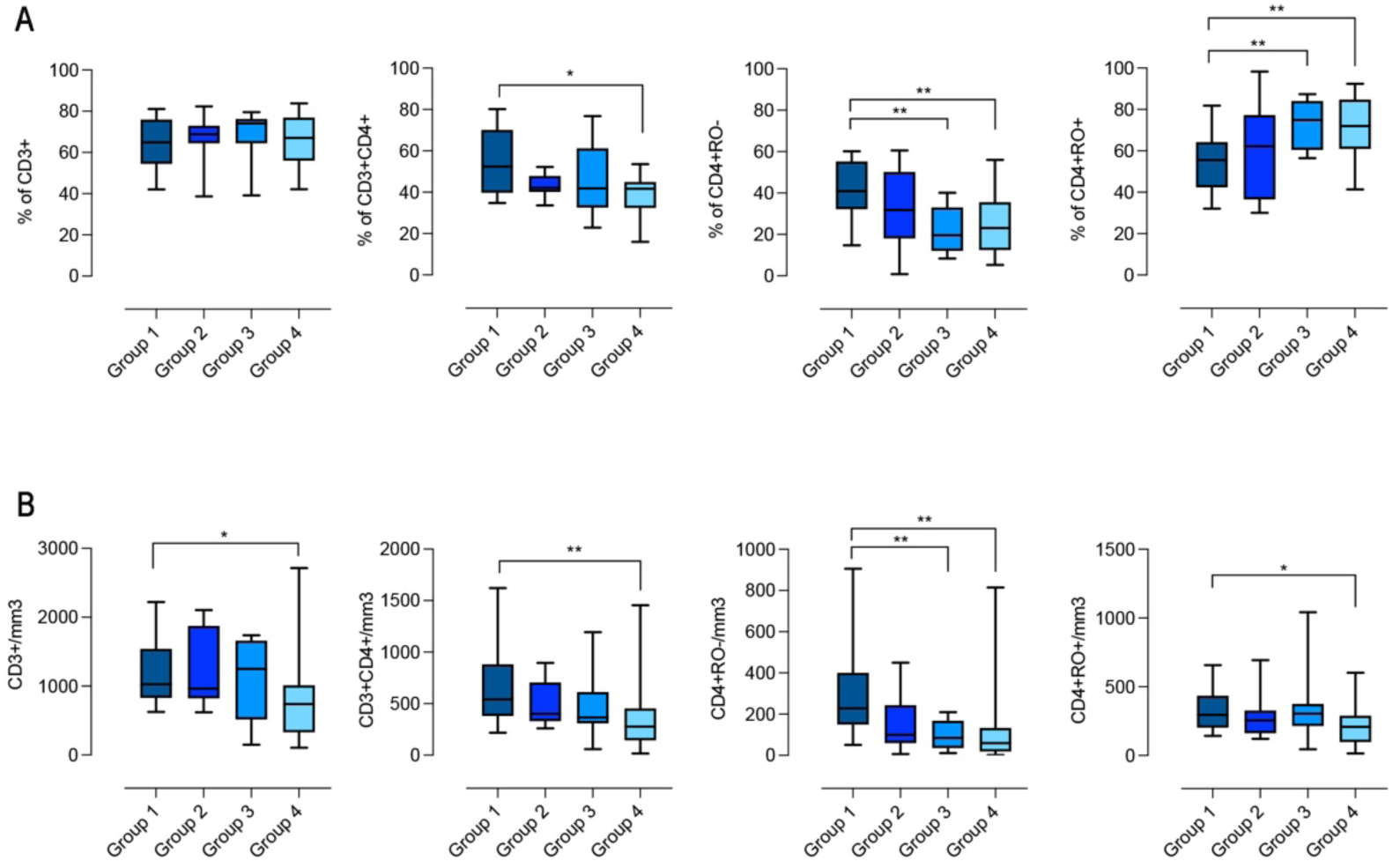
Frequency and absolute T-cell number in the 4 groups. (A-B) Bar plots depict the frequency (A) and the absolute number (B) of total T cells (CD3+), CD4+ T cells (CD3+CD4+), naive CD4+ (CD3+CD4+RO-) and memory CD4+ (CD3+CD4+RO+). Non-parametric Mann–Whitney t-test was used to evaluate statistical significance. Two-tailed P value significances are shown as * p<0.05, **p < 0.01.

**Supplementary Table 1.** List of 53 genes included in the IEIs panel.

| Gene |  |  |  |  |  |  |
| --- | --- | --- | --- | --- | --- | --- |
| ADA | CD19 | CR2 | IL2RA | NFKBIA | RAG1 | TNFRSF13C |
| AICDA | CD3G | CTLA4 | KRAS | NRAS | RAG2 | TTC7A |
| ATM | CD40 | DKC1 | LIG1 | OAS1 | SH2D1A | UNG |
| BLNK | CD40L | DCLRE1 | LRBA | PIK2CD | STAT1 | WAS |
| BTK | CD79A | FAS | MAGT1 | PIK3R1 | STAT3 | XIAP |
| CASP8 | CD79B | FASL | MS4A1 | PLCG2 | STX11 |  |
| CASP10 | CD81 | ICOS | NFKB1 | PRF1 | TCF3 |  |
| CD179B | CECR1 | IKZF1 | NFKB2 | PRKCD | TNFRSF13B |  |

**Supplementary Table 2.** Anti S1-IgG serum levels and frequencies of peripheral specific B-cells measure of d at the different study time in CVID patients naïve to SARS-CoV-2 infection

|  | Group 1(S1 IgG+/S++ MBC+) |  |  | Group 2 (S1 IgG+/S+ MBC+) |  |  | Group 3 (S1 IgG+/S+ MBC) |  |  | Group 4 (S1 IgG-/S+ MBC-) |  |  |
| --- | --- | --- | --- | --- | --- | --- | --- | --- | --- | --- | --- | --- |
|  | n=14 |  |  | n=7 |  |  | n=9 |  |  | n= 17 |  |  |
|  | Median | IQR |  | Median | IQR |  | Median | IQR |  | Median | IQR |  |
| S1 IgG post 3<sup>rd</sup> (OD ratio) | 33.8 | 24.47 | 48.80 | 3.87 | 1.46 | 15.62 | 2.76 | 2.07 | 14.52 | 0.25 | 0.16 | 0.41 |
| S1 IgG pre 4<sup>th</sup> (OD ratio) | 13.33 | 9.22 | 18.01 | 9.45 | 3.51 | 20.47 | 3.33 | 2.66 | 7.03 | 0.57 | 0.33 | 0.93 |
| S1 IgG post 4<sup>th</sup> (OD ratio) | 51.18 | 18.47 | 61.75 | 14.09 | 9.17 | 26.56 | 9.64 | 5.6 | 12.94 | 1.09 | 1.00 | 1.35 |
| S+ MBC post 3<sup>rd</sup> (%) | 0.26 | 0.22 | 0.33 | 0.13 | 0.12 | 0.22 | 0 | 0 | 0 | 0 | 0 | 0 |
| S+ MBC pre 4<sup>th</sup> (%) | 0.47 | 0.37 | 0.59 | 0.37 | 0.14 | 0.68 | 0 | 0 | 0 | 0 | 0 | 0 |
| S+MBC post 4<sup>th</sup> (%) | 0.50 | 0.39 | 0.79 | 0.33 | 0.18 | 0.67 | 0 | 0 | 0.37 | 0 | 0 | 0 |
| S++ MBC post 3<sup>rd</sup> (%) | 0.09 | 0.06 | 0.39 | 0 | 0 | 0 | 0 | 0 | 0 | 0 | 0 | 0 |
| S++ MBC pre 4<sup>th</sup> (%) | 0.11 | 0.06 | 0.37 | 0 | 0 | 0 | 0 | 0 | 0 | 0 | 0 | 0 |
| S++ MBC post 4<sup>th</sup> (%) | 0.44 | 0.22 | 0.55 | 0.02 | 0 | 0.13 | 0 | 0 | 0 | 0 | 0 | 0 |
| RBD+ post 3<sup>rd</sup> (%) | 10.1 | 7.4 | 19.5 | 0 | 0 | 0 | 0 | 0 | 0 | 0 | 0 | 0 |
| RBD+ pre 4<sup>th</sup> (%) | 15.8 | 4.7 | 24.6 | 0 | 0 | 1.5 | 0 | 0 | 0 | 0 | 0 | 0 |
| RBD+ post 4<sup>th</sup> (%) | 22.9 | 13.3 | 32.9 | 8.9 | 0 | 19.5 | 0 | 0 | 0 | 0 | 0 | 0 |

**Supplementary Table 3.**
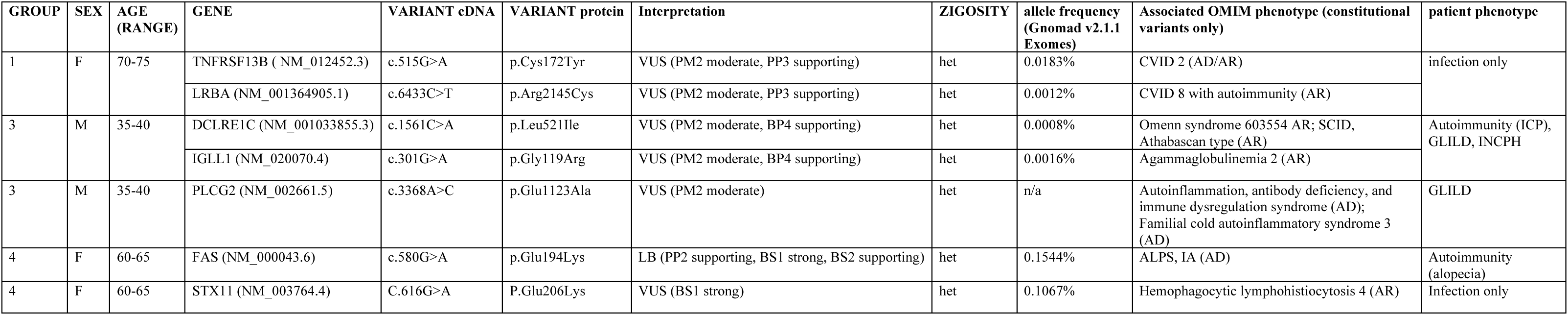
Polymorphism analysis

